# Lower Dynamic Cerebral Autoregulation following Acute Bout of Low-Volume High-Intensity Interval Exercise in Chronic Stroke Compared to Healthy Adults

**DOI:** 10.1101/2023.06.13.23291289

**Authors:** Alicen A. Whitaker, Stacey E. Aaron, Mark Chertoff, Patrice Brassard, Jake Buchanan, Katherine Nguyen, Eric D. Vidoni, Saniya Waghmare, Sarah M. Eickmeyer, Sandra A. Billinger

## Abstract

Fluctuating blood pressure during high-intensity interval exercise (HIIT) may challenge dynamic cerebral autoregulation (dCA), specifically post-stroke after an injury to the cerebrovasculature. We hypothesized dCA would be attenuated at rest and during a sit-to-stand, immediately following and 30 min after HIIT in individuals post-stroke compared to age- and sex-matched controls (CON). HIIT switched every minute between 70% and 10% estimated maximal watts for 10 min. Mean arterial pressure (MAP) and middle cerebral artery blood velocity (MCAv) were recorded. Resting dCA measured spontaneous fluctuations in MAP and MCAv via transfer function analysis. For sit-to-stand, time delay before an increase in cerebrovascular conductance index (CVCi = MCAv/MAP), rate of regulation, and %change in MCAv and MAP were measured. Twenty-two individuals post-stroke (age 60±12 yrs, 31±16 months) and twenty-four CON (age 60±13 yrs) completed the study. VLF gain (p=0.02, η^2^=0.18) and normalized gain (p=0.01, η^2^=0.43) had a group-by-time interaction, with CON improving immediately and 30 min after HIIT. Individuals post-stroke had impaired VLF phase (p=0.03, η^2^=0.22) immediately following HIIT compared to CON. We found no differences in the sit-to-stand. Our study showed lower resting dCA up to 30 min after HIIT in individuals post-stroke compared to CON while the sit-to-stand response was maintained.

## Introduction

Dynamic cerebral autoregulation (dCA), or the cerebral vasculature’s ability to react to rapid changes in mean arterial pressure (MAP),^1–3^ is impaired after stroke,^4–8^ At rest, dCA can be quantified using transfer function analysis using spontaneous fluctuations in MAP and cerebral artery blood velocity, to obtain gain (amplitude of cerebral blood velocity change for a given oscillation in MAP) and phase (difference in the timing of the MAP and cerebral blood velocity waveforms) metrics. dCA also responds to changes in MAP during everyday activities, such as standing up from a seated position.^3^ During a sit-to-stand, there is a transient drop in MAP, due to gravity and venous pooling. dCA then responds prior to the activation of the arterial baroreflex through an increase in cerebrovascular conductance index (CVCi).^3^ However, the effects of aerobic exercise on the dCA response in individuals post-stroke with impaired dCA are unknown.^9^

It is important to examine the ability of the cerebrovascular system to react to changes in MAP after aerobic exercise,^10, 11^ as dCA post-stroke may be less effective at buffering fluctuations in MAP, potentially leaving the brain less protected.^12^ In individuals with a pre-existing injury to the cerebrovascular system such as stroke, the dCA response should be an important factor in prescribing aerobic exercise and determining post-exercise recovery care.^12^ Specifically, one type of aerobic exercise, which may influence dCA and has gained in popularity for individuals post-stroke, is high-intensity interval exercise (HIIT).^13–15^

HIIT may challenge dCA due to the rapid fluctuations in MAP when switching between high-intensity exercise and recovery.^10, 11, 16^ Despite multiple narrative reviews suggesting HIIT may attenuate dCA to a larger degree in individuals with a cerebrovascular injury such as stroke compared to healthy adults,^10–12^ HIIT is starting to be implemented within stroke rehabilitation to improve motor outcomes.^13, 15, 17^ Therefore, we need to fill the gaps in knowledge on the effects of low-volume HIIT on cerebrovascular health and the dCA response in individuals with chronic stroke, specifically after exercise.^12^

While no studies have examined dCA after an acute bout of HIIT in individuals post-stroke,^9^ prior work in healthy adults reported dCA was less effective during repeated squat-stands following HIIT.^18^ During exercise recovery following HIIT, MAP decreases^16, 19^ which could challenge the dCA response, as dCA seems less effective at buffering decreases compared to increases in MAP at rest.^20^ During recovery following moderate to vigorous intensity continuous exercise in healthy adults, dCA at rest has been shown to be effected immediately following exercise and recover by 30 min.^21, 22^ However, the dCA response may take longer than 30 min to recover in individuals post-stroke potentially due to post-exercise hypotension and prolonged recovery of MAP.^12^ Recovery of end tidal carbon dioxide (P_ET_CO_2_) may also influence cerebral blood velocity following HIIT^23^ in individuals post-stroke compared to healthy adults^19^ and could also play a role in the dCA response during recovery.^16, 24, 25^

To address these critical gaps in knowledge, we characterized the dCA response using spontaneous fluctuations in MAP and middle cerebral artery blood velocity (MCAv) as well as during a single sit-to-stand following an acute bout of low-volume HIIT in individuals with chronic stroke compared to age- and sex-matched adult controls (CON). We hypothesized that individuals with chronic stroke would have attenuated dCA initially following HIIT as well as during a follow-up at 30 min after HIIT, compared to CON. We also hypothesized that P_ET_CO_2_ during recovery following HIIT would influence the dCA response.

## Material and Methods

The University of Kansas Medical Center Human Subjects Committee approved all study procedures and the Declaration of Helsinki and local statutory requirements were followed. This study was registered on clinicaltrials.gov (NCT04673994) and STROBE guidelines reported.

Our methods have been described previously.^16, 19^ Individuals with stroke were included within this study if they were: 1) between 40-85 years old, 2) treated for stroke 6 months to 5 years ago, 3) classified as inactive by performing <150 min of moderate intensity exercise per week, and 4) able to answer consenting questions and follow a 2-step command. Individuals within the CON group were matched to the sex and age ± 5 years of an individual post-stroke and also classified as inactive. Individuals were excluded from the study if: 1) stroke etiology was due to COVID-19, 2) they were unable to stand from a chair without physical assistance from another person, 3) they were unable to perform exercise on a recumbent stepper, 4) diagnosed with insulin-dependent diabetes, 5) diagnosed with another neurological disease, or 6) we were unable to find a transcranial Doppler ultrasound (TCD) signal on left or right side.

### Study Visit 1

All participants were informed of study procedures, benefits, and risks before providing voluntary written consent. We then collected participant demographics, current medications, prior medical history, and self-reported disability via the modified Rankin scale (mRS).^26^ Lower extremity function in individuals post-stroke was assessed using the Fugl Meyer Lower Extremity Subscale and the 5 Times Sit-to-Stand Assessment.^27, 28^ We then screened for an MCAv signal on both the left and right side using TCD probes (2-MHz, Multigon Industries Inc, Yonkers, New York). TCD signal acquisition was performed following standard depth, gain, and power.^29^ The position of the probes were recorded on *study visit 1* to ensure accurate signal acquisition on *study visit 2* when recordings were performed.

#### Submaximal Exercise Test

As previously reported,^16, 19^ the total body recumbent stepper (TBRS, T5XR NuStep, Inc. Ann Arbor, MI) submaximal exercise test was used to determine the estimated maximal watts (estimatedWatt_max_) for the acute HIIT bout.^30, 31^ If individuals were on a beta blocker medication, maximum HR was determined using the beta-blocker equation (164 – (0.7 * age)).

#### Exercise Familiarization

The low-volume HIIT exercise familiarization was performed at least 15 min after the TBRS submaximal exercise test or once HR and MAP returned to near baseline values. Based on previously published low-volume HIIT protocols in older adults and clinical populations, low-volume HIIT was dosed at 70% estimatedWatt_max_ and active recovery at 10% estimatedWatt_max_.^15, 32–37^ Participants practiced switching between high-intensity exercise and active recovery bouts every min for approximately 10 min.

### Study Visit 2

Following our laboratory protocols, the room was dimly lit and kept at a constant temperature (21 - 23°C).^16, 38^ All participants were asked to refrain from caffeine for 8 hours,^39^ food for 2 hours,^40^ vigorous exercise for 24 hours,^18^ and alcohol for 24 hours.^41^

#### Equipment Set-Up

Participants were seated on a recumbent stepper and fitted with the following equipment: 1) bilateral TCD probes were secured to an adjustable headset to ensure a stable position and angle to measure MCAv throughout testing, 2) a 5-lead electrocardiogram (ECG; Cardiocard, Nasiff Associates, Central Square, New York) to collect HR, 3) a left middle finger Finometer cuff (Finapres Medical Systems, Amsterdam, the Netherlands) to measure beat-to-beat MAP, 4) a right arm brachial automated sphygmomanometer with a microphone (Tango M2; Suntech, Morrisville, NC) for calibration of the Finometer, and 5) a nasal cannula attached to a capnograph (BCI Capnocheck Sleep 9004 Smiths Medical, Dublin, Ohio) for P_ET_CO_2_ monitoring. In individuals post-stroke with left arm spasticity, the Finometer was placed on the right arm.

#### Cerebral Autoregulation Measures

To quantify spontaneous dCA metrics, participants performed a 5-min seated rest recording 1) at BL, 2) immediately following HIIT and 3) 30 min after HIIT while capturing MCAv, MAP, HR, and P_ET_CO_2_ (**Figure 1**).

**Figure 1.**
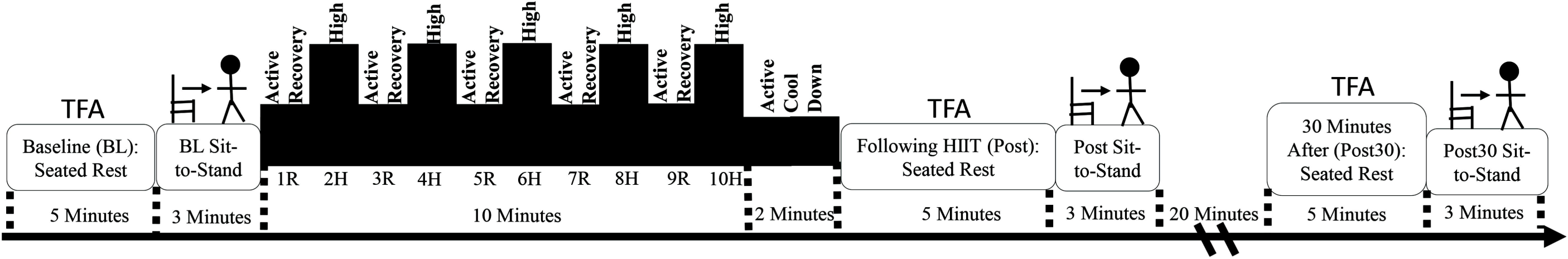
Protocol Measuring Dynamic Cerebral Autoregulation after an Acute Bout of HIIT. TFA = transfer function analysis (was conducted during seated rest). BL = baseline. R = active recovery. H = high-intensity. Post = Recovery immediately following HIIT. Post30 = Recovery at 30 min after HIIT.

Using a sit-to-stand procedure, we recorded MCAv, MAP, HR, and P_ET_CO_2_ and quantified dCA, based on our previously published methodology, 1) at BL, 2) following 5 min of passive recovery after HIIT, and 3) 35 min after HIIT.^42^ Participants placed the hand with the Finometer across their chest and a sling was used to secure the arm at heart-level during the sit-to-stand recording.^42^ Participants placed their feet flat on the ground, sat in an upright posture, and had a force-sensitive resistor (SEN-09376, Mouser Electronics Inc., Tx) to detect the exact moment of stance from the chair or the moment of arise-and-off (AO).^42^ The sit-to-stand recording lasted a total of 3 minutes (see Figure 1). Individuals began in a seated position for 1 min and then were given a 3-s countdown and asked to quickly transition to standing at 60-s and remained standing for 2 min for hemodynamic stability. Study personnel were standing next to the participants during the sit-to-stand transition. Initial orthostatic hypotension (within 15 s of standing) was determined by a drop of systolic blood pressure >40 mmHg and a drop in diastolic blood pressure >20 mmHg as well as symptoms of dizziness, blurry vision, nausea, or weakness with standing.^43–45^

#### Low-Volume HIIT Protocol

Repetitive 1-min intervals of high-intensity exercise (70% estimatedWatt_max_) separated by 1-min active recovery bouts (10% estimatedWatt_max_) were performed for 10 min.^16, 19^ The first minute of HIIT was an active recovery bout, to avoid a Valsalva Maneuver, which would cause a transient decrease in MCAv.^46^ Following HIIT, resistance and step rate were decreased for a 2-min cool-down.

#### Data Acquisition

Data was collected at 500 Hz via an analog-to-digital unit (NI-USB-6212, National Instruments) and custom-written code within MATLAB (v2014a, TheMathworks Inc, Natick, Massachusetts).^38^ Beat-to-beat data was processed offline using the QRS complex of the ECG.^38^ The left MCAv signals for CON was used for analysis. However, if left MCAv was not obtainable or had noise, the right MCAv signal was used.^38^ In individuals post-stroke, the ipsilesional hemisphere’s MCAv signal was used to compare to CON.

#### Transfer Function Analysis

Quantification of spontaneous TFA metrics followed the recommendations of the Cerebrovascular Research Network (CARNet).^47, 48^ The 5-min seated rest recordings were computed to cross spectral analysis via fast Fourier transform utilizing Welch’s method, 100-s Hanning windows, and 50% superposition. Waveforms of MAP and MCAv were analyzed via TFA and described through gain (amplitude), phase (shift in radians), and coherence (association). Due to dCA acting as a high-pass filter, measures of gain, phase, coherence, and power spectral density (PSD) of the integrate area were reported within very low frequency (0.02 – 0.07 Hz) and low frequency (0.07 – 0.2Hz) bands. Following CARNet recommendations, the critical threshold of coherence was set at 0.34.^47^ Coherence determined whether estimates of gain and phase were reliable and could be compared across time.^47^ We visually inspected our data for phase wrap-around and if detected these data points were removed.^47^ Attenuated dCA was interpreted as higher gain (greater differences in amplitude) and lower phase (smaller shift) following HIIT compared to baseline.

#### Sit-to-Stand Analysis

Metrics of dCA included the time delay (TD) before the onset of the regulatory response, rate of regulation (ROR), percent change in MCAv (%ΔMCAv), percent change in MAP (%ΔMAP), and the ratio of %ΔMCAv/%ΔMAP. To measure the TD before the onset of the regulatory response, beat-to-beat CVCi was calculated (CVCi = MCAv/MAP). Based on previously published methods,^2, 42^ 2 trained researchers selected the physiological heart beat when CVCi began to increase (without immediate transient reduction).^2^ The TD before the onset of the regulatory response was calculated as the time in s from AO until the continuous increase in CVCi. The ROR was used to determine the efficiency of the regulatory response, by calculating the slope of the 3 heart-beats following TD (ROR = ΔCVCi/Δt/ΔMAP).^2^ The ROR is taken within phase 1 of the sit-to-stand response (1-7 s after standing), prior to the onset of the arterial baroreflex response.^3, 43^ The %ΔMCAv and %ΔMAP were calculated from the seated baseline (15 s prior to AO) to nadir (the lowest value of MCAv and MAP after standing).

%Δ = ((seated baseline – nadir) / seated baseline) * 100

An attenuated dCA response was interpreted as a longer TD before the onset of the regulatory response and smaller ROR, greater %ΔMCAv and a higher ratio of %ΔMCAv/%ΔMAP.^2, 43^

#### Statistical Analysis

A Shapiro-Wilk test was used to verify the assumptions of normality and a priori α was set at 0.05. Participant characteristics were compared using an independent t-tests for continuous variables or the Mann-Whitney U tests for non-normally distributed continuous variables.

Categorical variables were compared using a Chi-square. Baseline differences between groups were compared using an independent t-tests. The primary aim of this study was to examine dCA during spontaneous fluctuations at rest and during a sit-to-stand: 1) at BL, 2) following HIIT, and 3) at a follow up 30 min after HIIT in individuals post-stroke compared to CON. Mixed-model ANOVAs were used to determine group, time, and group by time interaction effects. Post-hoc comparisons used one-sided t-tests. As a sensitivity analysis, we controlled for P_ET_CO_2_ after HIIT within the mixed model ANOVAs.

## Results

Sixty individuals were enrolled within the study. Forty-six individuals were included in the primary analysis: individuals post-stroke (n = 22) and CON (n = 24), shown in **Figure 2**.

**Figure 2.**
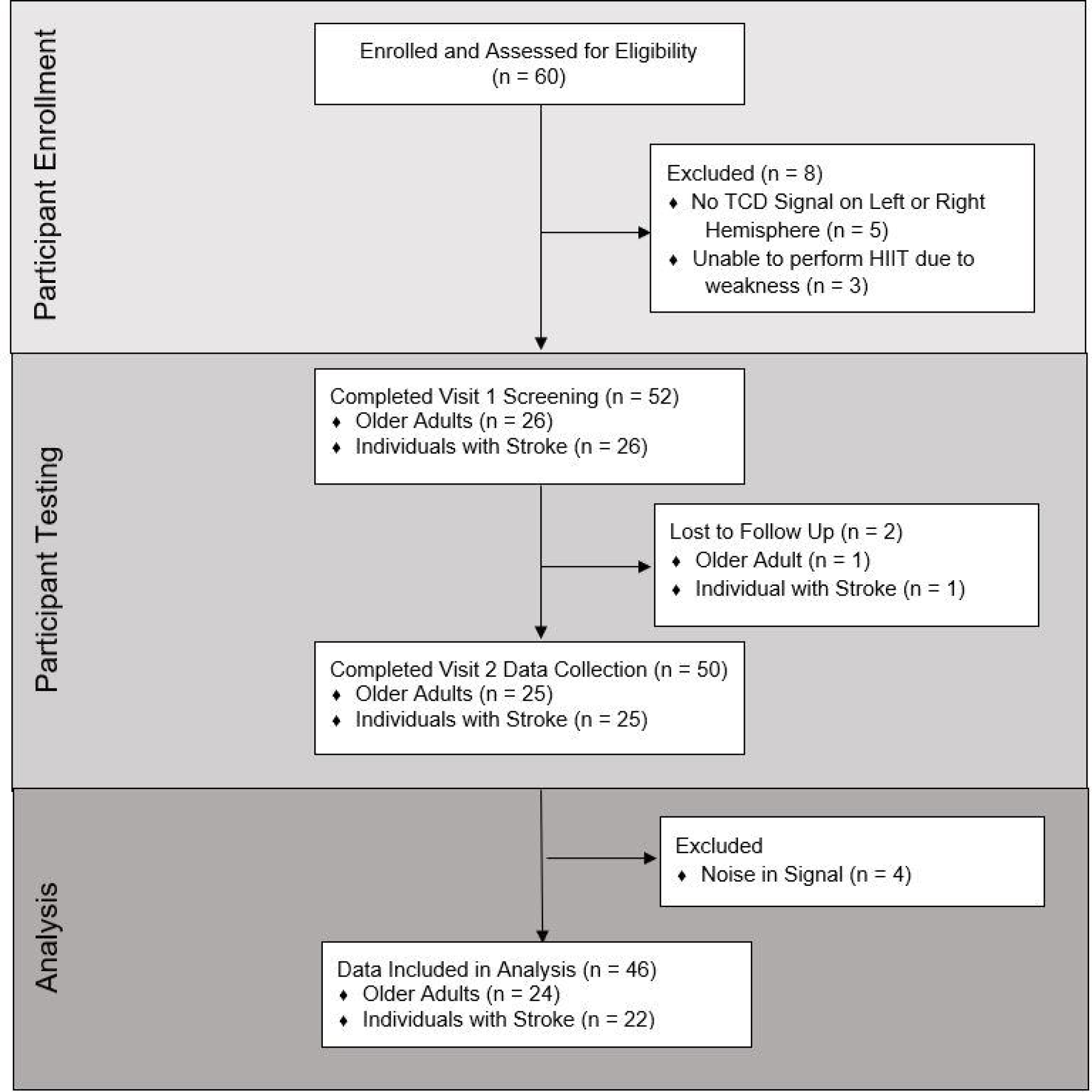
Flow Diagram.

Participant characteristics are described in **Table 1** and have been published in part within our previous work.^16, 19^ Briefly, individuals post-stroke had a significantly slower 5x sit-to-stand, lower average workload at high-intensity, and lower workload during active recovery compared to CON. Individuals post-stroke had slight-to-moderate disability as shown by the mRS and lower extremity Fugl-Meyer sub score.^49^

**Table 1.** Participant Characteristics.

|  | **Individuals**<br>**Post-Stroke**<br>**(n = 22)** | **CON**<br>**(n = 24)** | **p-value** |
| --- | --- | --- | --- |
| **Age<sup>†</sup> (years)** | 60 ± 12 | 60 ± 13 | 0.80 |
| **Female n (%)** | 9 (41%) | 8 (33%) | 0.76 |
| **BMI (kg/m<sup>2</sup>)** | 30.7 ± 6.0 | 28.9 ± 6.6 | 0.34 |
| **Race n (%)** |  |  | 0.34 |
| Asian | 0 | 1 (4%) |  |
| Black/African American | 4 (18%) | 2 (8%) |  |
| White/Caucasian | 17 (77%) | 21 (88%) |  |
| White & Native American | 1 (5%) | 0 |  |
| **Blood Pressure Medications** |  |  |  |
| Beta Blocker | 9 (41%) | 1 (4%) | **0.004\*** |
| ACE Inhibitor | 6 (27%) | 2 (8%) | 0.13 |
| Angiotensin II Receptor<br>Blocker | 6 (27%) | 5 (21%) | 0.73 |
| Calcium Channel Blocker | 8 (36%) | 3 (13%) | 0.09 |
| Diuretic | 2 (9%) | 3 (13%) | 1.00 |
| Vasodilator | 1 (5%) | 0 | 0.48 |
| **5x Sit-to-Stand<sup>†</sup> (s)** | 21.9 ± 13.5 | 8.6 ± 2.7 | **<0.001\*** |
| **TBRS estimated $\dot{V}\dot{O}_{2\max}$**<br>($\text{ml}\cdot\text{kg}^{-1}\cdot\text{min}^{-1}$) | $29.2 \pm 9.4$ | $32.3 \pm 10.2$ | 0.30 |
| **Workload during High Intensity<sup>†</sup>**<br>(Watts) | $79 \pm 27$ | $117 \pm 36$ | <0.001* |
| **Workload during Active Recovery<sup>†</sup>**<br>(Watts) | $16 \pm 2$ | $18 \pm 4$ | 0.04* |
| **<u>Stroke Characteristics</u>** |  |  |  |
| **Months Post-Stroke** | $31 \pm 16$ | | |
| **Right-Sided Stroke n (%)** | 11 (50%) |  |  |
| **Ischemic Stroke n (%)** | 18 (82%) |  |  |
| **MCA Stroke n (%)** | 12 (55%) |  |  |
| **mRS n (%)** |  |  |  |
| No Significant Disability (1) | 9 (41%) |  |  |
| Slight Disability (2) | 6 (27%) |  |  |
| Moderate Disability (3) | 7 (32%) |  |  |
| **Fugl-Meyer Lower Extremity Score** | $23 \pm 6$ | | |
| Means (Standard Deviations). *Significantly different between groups. † = Non-normally distributed. BMI = body mass index. TBRS = Total Body Recumbent Stepper Submaximal Exercise Test. $\dot{V}\dot{O}_{2\max}$ = maximal oxygen consumption. $\text{ml}\cdot\text{kg}^{-1}\cdot\text{min}^{-1}$ = milliliters of oxygen per kilogram of body weight per minute. estimatedWatt<sub>max</sub> = estimated maximal watts. mRS = modified rankin scale. MOCA = montreal cognitive assessment. | | | |

### Dynamic Cerebral Autoregulation during Spontaneous Fluctuations in MAP and MCAv

Twenty-two individuals had complete TFA data sets across all 3 timepoints, shown in **Table 2**. The reduction in TFA sample size is common and occurs due to reduced coherence within the VLF range,^47^ which could be attributed to non-linearity.^50^ At BL, there was no significant differences between groups in all measures. MAP PSD and MCAv PSD within the VLF and LF range were also not different between groups or across time.

**Table 2.**
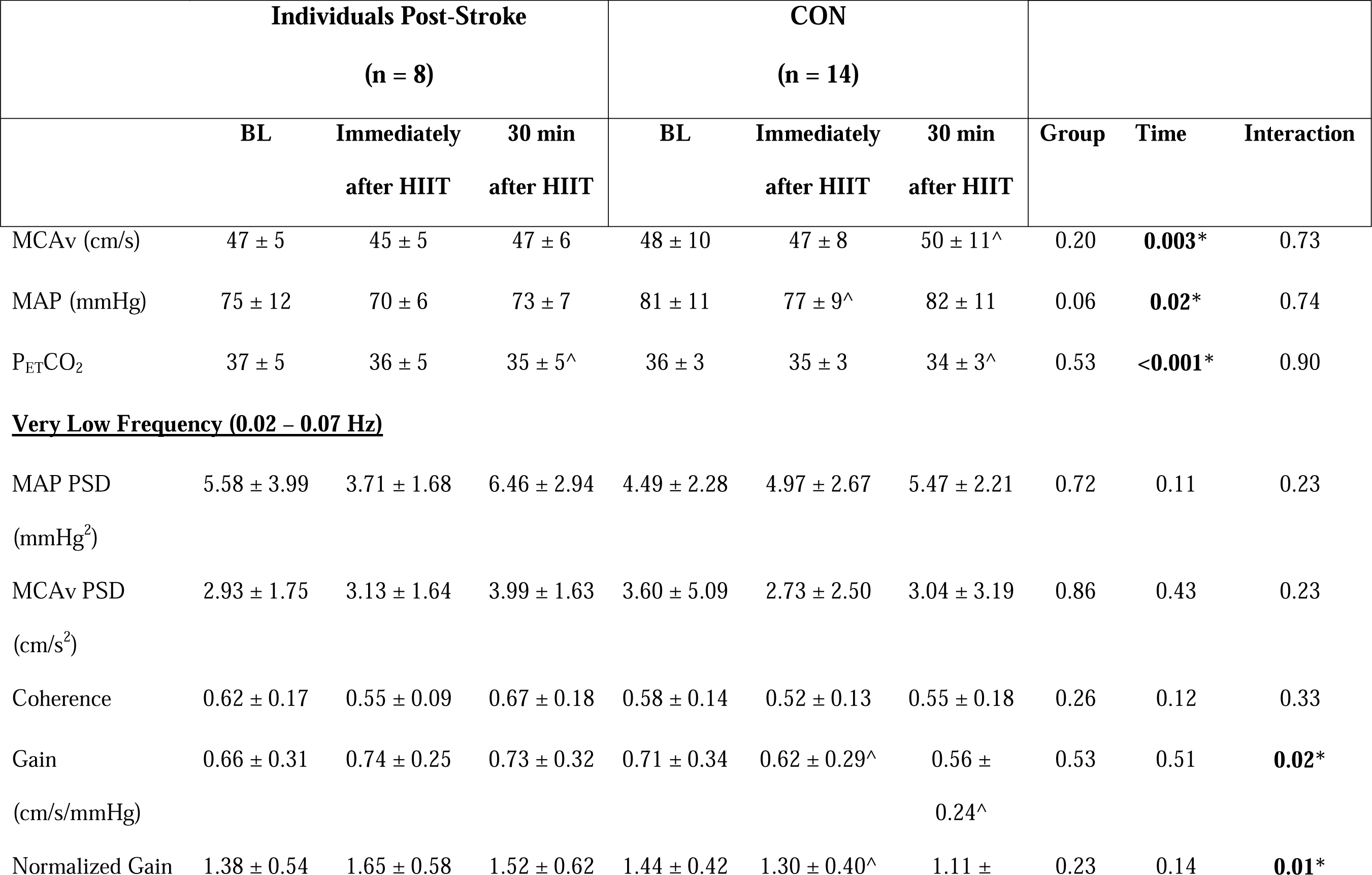
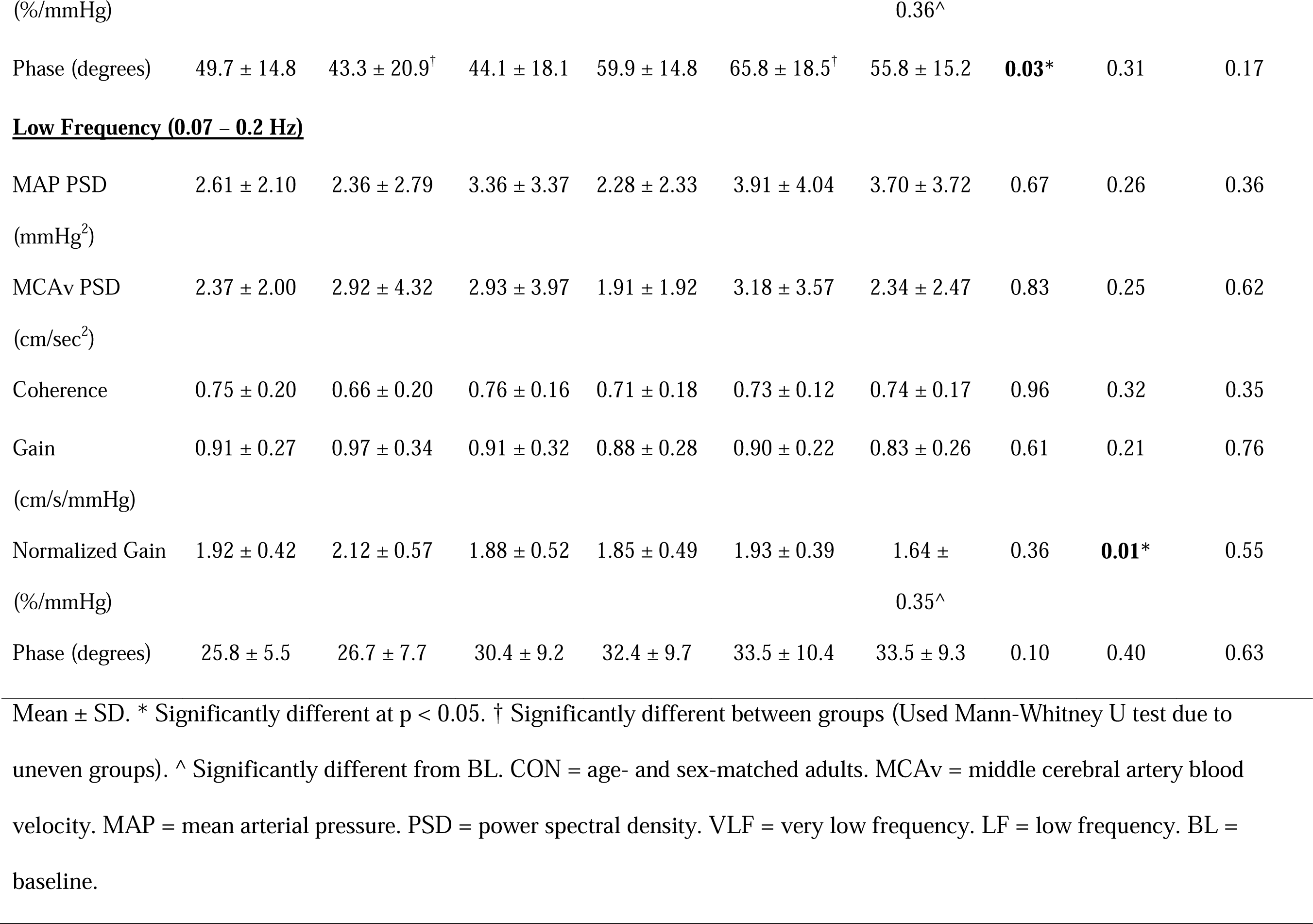
Transfer Function Analysis of Spontaneous Fluctuations at Rest.

Our primary analysis showed a significant group by time cross-over interaction effect for VLF gain (η^2^ = 0.18) and VLF normalized gain (η^2^ = 0.43), with the CON group having decreased VLF gain and normalized gain immediately following HIIT and 30 min after HIIT compared to BL. While average VLF gain and normalized gain in individuals post-stroke changed in the opposite direction of CON following HIIT, creating a cross-over interaction, there was no significant within-group differences across time. There was also a significant group effect for VLF phase (η^2^ = 0.22), with individuals post-stroke having lower VLF phase compared to CON immediately following HIIT (p = 0.01), and returned to being not significantly different between groups at 30 min after HIIT. Finally, there was a linear time effect for LF normalized gain, where CON individuals had reduced normalized gain at 30 min following HIIT compared to BL (p = 0.03).

Our sensitivity analysis showed that when controlling for reductions in P_ET_CO_2_ after HIIT, the group by time interaction remained for VLF gain (p = 0.01) and VLF normalized gain (p = 0.004). The group effect for VLF phase (p = 0.046) also remained when controlling for P_ET_CO_2_. However, the time effect for LF normalized gain (p = 0.56) was no longer significant when controlling for P_ET_CO_2_. The transfer function analysis across the frequency ranges (0.02 – 5 Hz) in individuals with chronic stroke compared to CON are shown in **Figure 3**.

**Figure 3.**
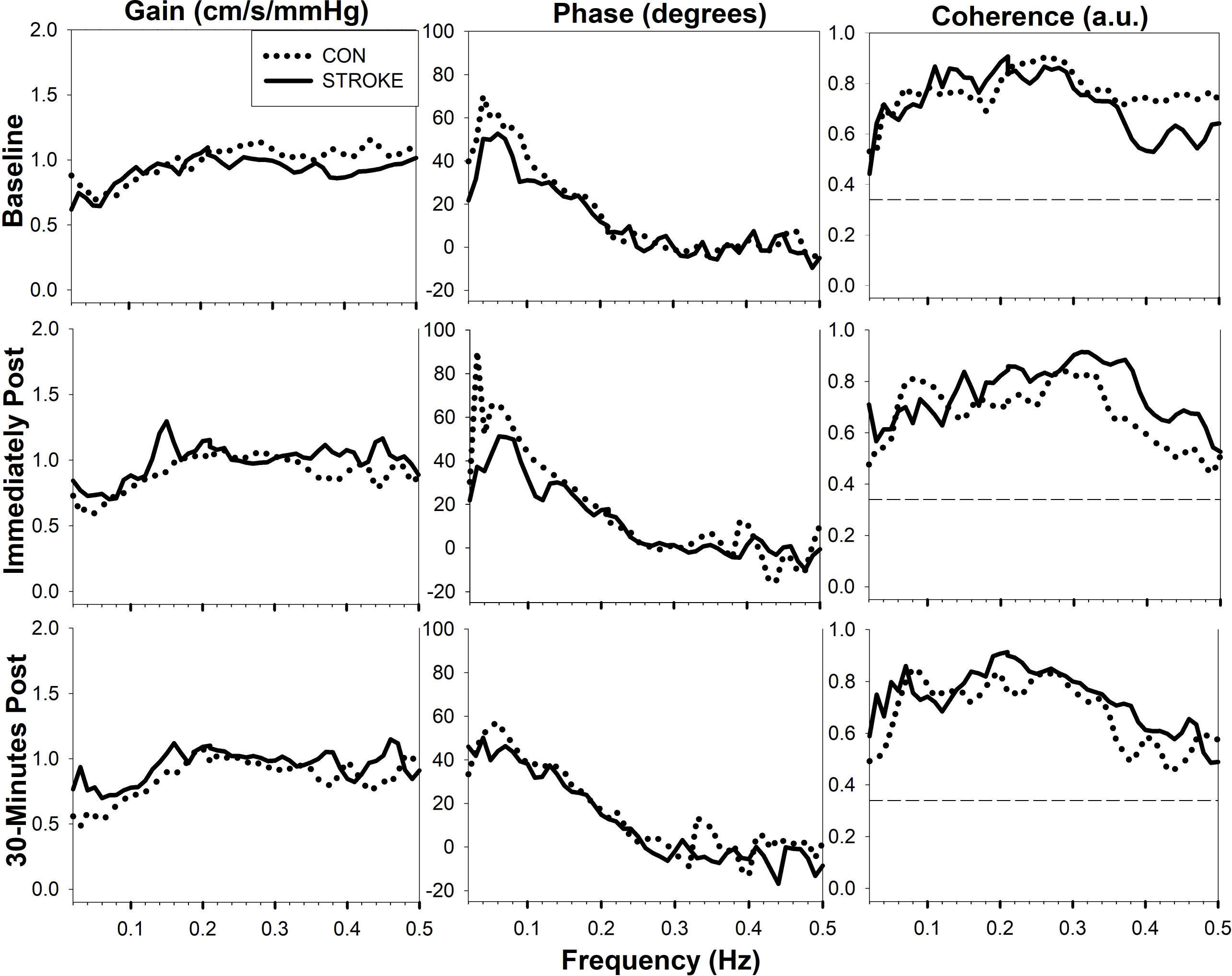
Transfer Function Analysis in Individual Post-Stroke compared to CON following HIIT. Solid black line = individuals with chronic stroke. Dotted Line = age- and sex-matched adult controls (CON). Hz = hertz.

### Sit-to-Stand Measures of Dynamic Cerebral Autoregulation

There were no significant group or time effects on the sit-to-stand measures of dCA following HIIT in individuals post-stroke compared to CON, shown in **Table 3**. When controlling for seated P_ET_CO_2_ after HIIT, there were still no group or time effects for TD onset, ROR, %ΔMCAv, %ΔMAP, or %ΔMCAv/%ΔMAP.

**Table 3.** Sit-to-Stand Dynamic Cerebral Autoregulation Metrics.

|  | **Individuals Post-Stroke** |  |  | **CON** |  |  |  |  |  |
| --- | --- | --- | --- | --- | --- | --- | --- | --- | --- |
|  | **(n = 22)** |  |  | **(n = 23)** |  |  |  |  |  |
|  | **BL** | **Following**<br>**HIIT** | **35 min**<br>**after HIIT** | **BL** | **Following**<br>**HIIT** | **35 min**<br>**after HIIT** | **Group** | **Time** | **Interaction** |
| **Seated MCAv (cm/s)** | 42 ± 11 | 42 ± 11 | 42 ± 11 | 50 ± 15 | 49 ± 14 | 49 ± 13 | 0.052 | 0.07 | 0.66 |
| **Seated MAP (mmHg)** | 84 ± 12 | 82 ± 10 | 82 ± 8 | 82 ± 11 | 86 ± 11 | 85 ± 11 | 0.52 | 0.49 | 0.08 |
| **Seated P<sub>ET</sub>CO<sub>2</sub> (mmHg)** | 36 ± 5 | 34 ± 6<sup>^</sup> | 35 ± 4 | 36 ± 4 | 34 ± 4<sup>^</sup> | 34 ± 4<sup>^</sup> | 0.71 | **<0.001\*** | 0.31 |
| **Standing MCAv (cm/s)** | 41 ± 11 | 39 ± 11 | 41 ± 11 | 48 ± 14 | 47 ± 13 | 48 ± 13 | 0.60 | 0.001 | 0.53 |
| **Standing MAP (mmHg)** | 84 ± 14 | 84 ± 14 | 84 ± 13 | 83 ± 13 | 85 ± 12 | 87 ± 12 | 0.74 | 0.32 | 0.26 |
| **Standing P<sub>ET</sub>CO<sub>2</sub> (mmHg)** | 36 ± 4 | 34 ± 4<sup>^</sup> | 35 ± 4<sup>^</sup> | 35 ± 3 | 33 ± 4<sup>^</sup> | 33 ± 4<sup>^</sup> | 0.27 | **<0.001\*** | 0.51 |
| **TD onset** | 2.99 ± 2.05 | 2.88 ± | 2.66 ± 2.00 | 3.00 ± | 2.83 ± | 2.99 ± 2.77 | 0.83 | 0.90 | 0.87 |
|  |  | 1.97 |  | 2.14 | 1.86 |  |  |  |  |
| **ROR** | 0.005 ± | 0.026 ± | 0.005 ± | 0.003 ± | 0.003 ± | 0.003 ± | 0.11 | 0.21 | 0.21 |
|  | 0.009 | 0.080 | 0.006 | 0.003 | 0.003 | 0.003 |  |  |  |
| **%ΔMCAv** | 12.1% ± | 16.9% ± | 14.9% ± | 15.3% ± | 15.9% ± | 16.0% ± | 0.66 | 0.17 | 0.32 |
|  | 10.0% | 8.2% | 9.7% | 10.1% | 11.6% | 10.1% |  |  |  |
| **%ΔMAP** | 16.5% ± | 18.0% ± | 15.5% ± | 18.7% ± | 19.8% ± | 18.5% ± | 0.35 | 0.22 | 0.88 |
|  | 8.5% | 7.7% | 9.9% | 8.9% | 10.8% | 9.6% |  |  |  |
| **%ΔMCAv/%ΔMAP** | 0.64 ± 0.61 | 1.06 ± | 0.87 ± 1.13 | 0.77 ± | 0.80 ± | 0.85 ± 0.50 | 0.80 | 0.19 | 0.30 |
|  |  | 1.12 |  | 0.47 | 0.63 |  |  |  |  |

Mean ± SD. \* Significantly different at p < 0.05. † Significantly different between groups. ^Significantly different than BL.
CON = age- and sex-matched adults. BL = baseline. TD = time delay. ROR = rate of regulation. %Δ = percent change from baseline to nadir upon standing. MCAv = middle cerebral artery blood velocity. MAP = mean arterial pressure.

**Figure 4** shows the CVCi response during the sit-to-stand at BL in individuals post-stroke compared to CON. Monitoring of beat-to-beat MAP during the sit-to-stand allows for the detection of initial orthostatic hypotension, defined by the literature as a decrease in systolic blood pressure >40 mmHg and/or diastolic blood pressure >20mmHg within 15 s after standing as well as symptoms of dizziness or syncope.^45^ These values differ from the clinical thresholds of delayed orthostatic hypotension (systolic >20mmHg and/or diastolic >10mmHg) taken after 3-min of standing.^51^ We report that no individuals met the criteria of initial orthostatic hypotension (having both decreases in blood pressure and symptoms of cerebral hypoperfusion)^45^ during our study. However, following HIIT, three CON individuals met the criteria for initial drops in blood pressure upon standing but reported no symptoms of dizziness. At 35-min after HIIT, 2 different CON individuals met the criteria for an initial drop in blood pressure upon standing. While no individuals post-stroke met the blood pressure thresholds of drops of >40mmHg systolic or diastolic >20mmHg upon standing, one individual post-stroke reported slight dizziness upon standing at baseline (lasting < 3 s) and another individual post-stroke reported slight dizziness upon standing (lasting <3 s) following HIIT.

**Figure 4.**
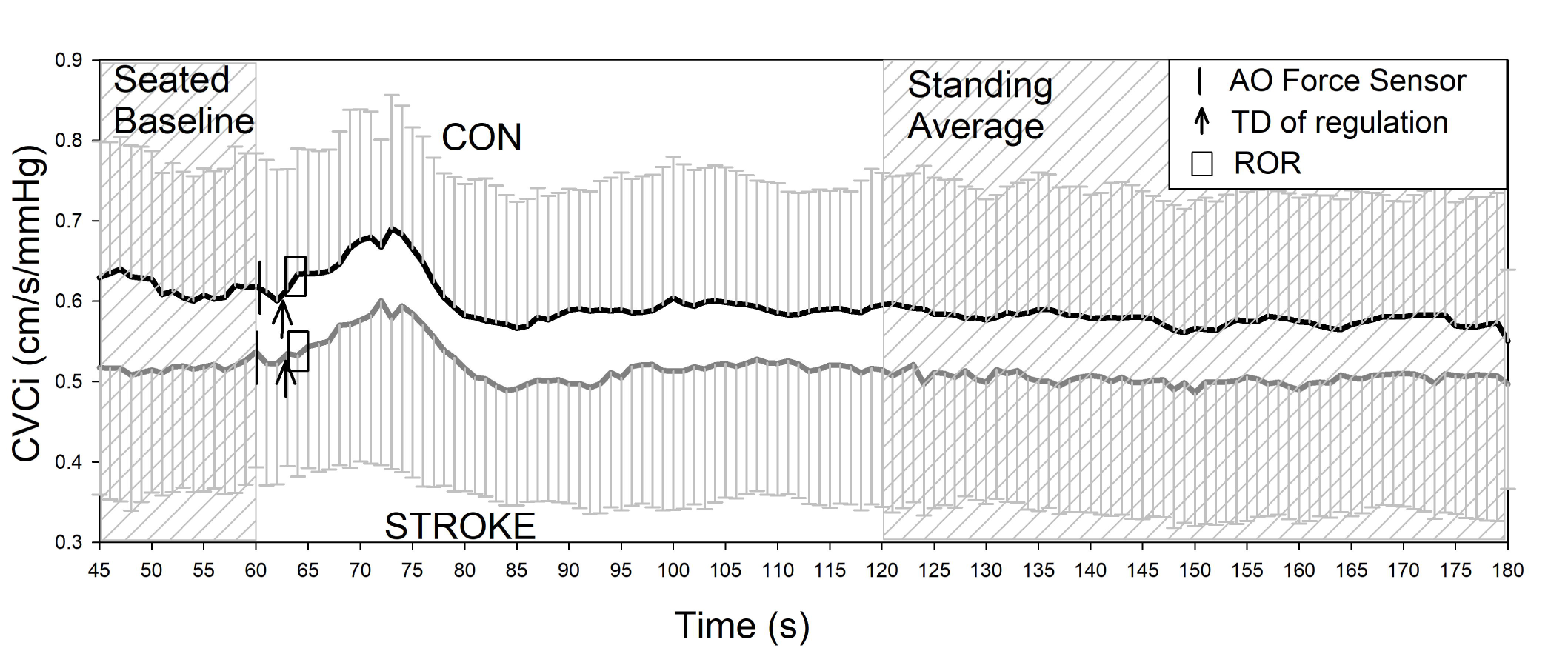
Baseline Sit-to-Stand Measure of Dynamic Cerebral Autoregulation. Mean and standard deviation lines. Individuals post-stroke (n = 22) = Grey line. Age- and sex-matched controls (CON, n = 23) = Black line. CVCi = cerebrovascular conductance index (MCAv/MAP). Horizontal black line = marks the moment of arise-and-off (AO) detected by the custom force sensor as the moment individuals stood up. Arrow = Time delay (TD) of the regulation response before an increase in CVCi. Square box = slope of the rate of regulation (ROR). Resampled to 1 Hz for group averages.

## Discussion

Our findings collectively show that while dCA was not different at baseline between individuals with chronic stroke and their age- and sex-matched peers, dCA was lower during spontaneous fluctuations in MAP and MCAv immediately following HIIT. While various reviews have outlined the importance of examining the dCA response to HIIT in individuals post-stroke,^10–12^ our study is the first to show: 1) individuals post-stroke had lower VLF phase immediately following HIIT compared to CON, and 2) VLF gain and normalized gain had a cross-over interaction effect in which CON individuals had a significant decrease in gain immediately following HIIT and 30 min after HIIT compared to BL while TFA gain in individuals post-stroke increased. Reductions in P_ET_CO_2_ following HIIT only accounted for the significant difference in LF normalized gain. Interestingly, while dCA was lower following HIIT during spontaneous MAP and MCAv fluctuations, our study did not find differences in the sit-to-stand dCA response between individuals post-stroke and CON, even when controlling for P_ET_CO_2_. Therefore, dCA in individuals post-stroke may maintain the ability to respond to a single drop in MAP upon standing, after 6 min of seated rest recovery following HIIT.

### Spontaneous Dynamic Cerebral Autoregulation

While previous systematic reviews have reported impaired resting dCA in individuals post-stroke compared to controls,^52^ our results did not show a difference between groups at baseline. Most of the previous studies reporting impaired dCA in individuals post-stroke were conducted during the acute stage (1-7 days post) and subacute stage (7 days – 6 months) post-stroke.^53–55^ To our knowledge, only 1 other study has examined dCA in the chronic stage (≥ 6 months) post-stroke and reported decreased resting phase (within 0.06 – 0.12 Hz) at exactly 6-months post-stroke compared to healthy controls.^56^ However, our study found no difference in resting BL dCA between individuals 31 ± 16 months post-stroke and their age- and sex-matched peers. This could be due to differences in supine vs. seated rest recordings, TFA frequency ranges utilized, or the chronicity of stroke. It is possible that dCA at rest during spontaneous blood pressure oscillations may continue to recover within the chronic stage post-stroke.^52^

To show differences in dCA between individuals with chronic stroke and healthy adults, the cerebrovascular system may need to be hemodynamically challenged, such as with an acute bout of HIIT. We show that immediately following HIIT, dCA during spontaneous MAP and MCAv fluctuations was lower in individuals post-stroke compared to CON, with lower VLF phase and increased VLF gain and normalized gain. Lower VLF phase immediately following HIIT in individuals post-stroke compared to CON can be interpreted as dCA being less efficient at buffering the cerebrovascular system from changes in peripheral blood pressure.^57^ The cross-over interaction in VLF gain and normalized gain shows that while CON individuals had significant improvements in the ability of the cerebrovasculature to dampen the amplitude changes in MAP fluctuations^57^ immediately following HIIT and 30 min after HIIT, individuals post-stroke had a reduction in the cerebral vessels’ dampening ability. Even when controlling for changes in P_ET_CO_2_, the differences in dCA between groups during recovery were maintained within the VLF range.

Lower dCA in individuals post-stroke, quantified via TFA immediately following HIIT, could be due to dCA’s sensitivity to decreasing MAP.^19, 58, 59^ Previous studies have shown that dCA at rest^20^ and during repetitive squat-stands^58–60^ was less efficient at regulating decreases in MAP in comparison with MAP increases, and HIIT may exacerbate this directional sensitivity in healthy adults.^61^ It is unknown whether individuals post-stroke have lesser dCA sensitivity to decreasing MAP compared to CON, however, this could potentially explain the differences between groups found immediately following HIIT. However, when forcing blood pressure to decrease using a single sit-to-stand following HIIT, we did not find any differences in the cerebral pressure-flow relationship in individuals post-stroke. Previous studies have also shown that blood pressure medications such as beta-blockers and calcium channel blockers may attenuate dCA.^62–65^ Our study reports individuals post-stroke had greater beta-blocker use and trended toward greater calcium channel blocker use compared to CON. While we did not directly measure the effects of blood pressure medications on dCA, they could have also contributed to the lower dCA response in individuals post-stroke following HIIT.

### Sit-to-Stand Dynamic Cerebral Autoregulation Response

There were no differences between individuals post-stroke and CON in the sit-to-stand dCA response at BL and after HIIT. We did not find a significant change in the TD before the onset of dCA or the ROR response in individual post-stroke following HIIT compared to CON. Therefore, the temporal response of increasing CVCi during a sit-to-stand was maintained following HIIT in individuals post-stroke. A previous study in healthy adults has shown that the initial dCA response during a sit-to-stand (occurring 2 ± 2 heartbeats after standing) may be due in-large to compliance of the cerebrovasculature, which occurs prior to cerebral vasodilation (occurring 11 ± 3 heartbeats after standing).^66^ While not measured within the study, cerebrovascular compliance may play a role in the preserved dCA sit-to-stand response following HIIT in individuals post-stroke and aging adults. We also did not show a significant difference in %ΔMCAv/%ΔMAP following HIIT. These findings may suggest that the cerebrovascular system in individuals post-stroke did not become more pressure passive following HIIT, and changes in MAP when standing up from a seated position resulted in similar concordant changes in MCAv.^67^ However, when interpreting the dCA sit-to-stand findings, one must also take into account that the participants had a notable amount of seated recovery (∼6 min) before the dCA sit-to-stand measure. Therefore, future studies should examine the sit-to-stand response immediately following aerobic exercise, without seated rest, to determine stability of the dCA response and how long someone should wait before standing up.

### Clinical Importance

Within the clinic, traditionally only peripheral blood pressure is used to determine hemodynamic stability during exercise recovery. However, our study shows the need to also examine the cerebrovascular system in individuals post-stroke. While the clinical implications for a lower dCA response during passive recovery immediately following HIIT are unknown, clinicians may want to continue practicing the recommended post-exercise precautions within the first 5 minutes following HIIT.^12^ When waiting ≥ 6 min after HIIT to stand up from the seated position, individuals post-stroke did not differ in the dCA response compared to BL or CON. However, two individuals post-stroke did report slight dizziness upon standing, despite not meeting the peripheral blood pressure criteria for initial orthostatic hypotension. Symptomatic thresholds associated with drops in MCAv during sit-to-stands following exercise should be established^68^ in individuals post-stroke to better understand the cerebrovascular response rather than making inferences of “hypoperfusion” from peripheral blood pressure.

### Methodological Considerations

The results of this study cannot be generalized to the entire population of individuals post-stroke, such as individuals with more severe impairments. Our findings within the chronic stage of stroke also cannot be generalized to individuals within the acute and subacute stages, where dCA has been shown to be impaired at baseline.^52^ While TCD is currently the best method to capture beat-to-beat MCAv for TFA, one must assume a constant MCA diameter. While we could not directly measure MCA diameter during the study, a previous study using 4D Flow Magnetic Resonance Imaging in older adults showed no significant change in diameter during an 8-mmHg change in P_ET_CO_2_.^69^ Our sample size to quantify TFA metrics was also reduced due to low coherence, potentially due to non-linearity of spontaneous fluctuations.^50^ Although we followed the CARNet recommendation of a 5-min recording,^47^ to increase coherence via non-linear modeling requires longer recordings (∼40 minutes).^50^ Measuring the dCA response via a single sit-to-stand was chosen due to its real-life applicability as well as accounting for neuromuscular fatigue after HIIT in individuals post-stroke. While our sit-to-stand methodology accounted for the exact time in which individuals stood from the chair by using a force sensor, we did not use an accelerometer to account for the speed of standing. During the sit-to-stand, we were also unable to account for other potential confounding factors including neurovascular coupling, sympathetic activity, and cardiac output. Since accumulating evidence also suggests directional sensitivity of the cerebral pressure-flow relationships,^20, 58–61, 70, 71^ further research quantifying dCA while considering the direction of MAP changes will be necessary to better understand whether the reported attenuated dCA after HIIT is in response to reductions in MAP, increases in MAP, or both.

## Conclusion

We show that dCA at rest during spontaneous fluctuations in MAP and MCAv was lower following HIIT in individuals post-stroke compared to CON. However, the dCA response during a sit-to-stand following HIIT (after ∼6 min seated rest) did not show a significant difference compared to CON. These findings provide the groundwork for further characterization of the dCA response following aerobic exercise of varying intensities in individuals across the stages of stroke recovery. The cerebral autoregulation response to aerobic exercise and recovery is “an essential physiological consideration to protect the brain when progressing exercise intensity [post-stroke]”.^12^ Future research should develop therapeutic strategies to improve dCA post-exercise in individuals post-stroke.

## Data Availability

All data produced in the present study are available upon request to the authors.

## Acknowledgements

Thank you, Andrew Geise, Kailee Carter, Katelyn Struckle, and Emily Hazen for data collection.

## Author Contribution Statement

AW, MC, EV, SE, and SB, conceived and designed research; AW, SA, JB, KN, and SW, performed experiments; AW, analyzed data; AW, SA, MC, PB, EV, and SB interpreted results of experiments; AW, PB, SB drafted manuscript; AW, SA, JB, KN, MC, PB, EV, SW, SE, and SB edited and revised manuscript; AW, SA, JB, KN, MC, PB, EV, SW, SE, and SB approved final version of manuscript.

